# The role of asymptomatic and pre-symptomatic infection in SARS-CoV-2 transmission – a living systematic review

**DOI:** 10.1101/2020.09.01.20135194

**Authors:** Xueting Qiu, Ali Ihsan Nergiz, Alberto Enrico Maraolo, Isaac I. Bogoch, Nicola Low, Muge Cevik

**Author notes:** **Author of correspondence**: Name: Dr Muge Cevik, Address: Division of Infection and Global Health Research, School of Medicine, University of St Andrews, Fife, Scotland, KY16 9TF.

## Abstract

**Background:** Reports suggest that asymptomatic individuals (those with no symptoms at all throughout infection) with severe acute respiratory syndrome coronavirus 2 (SARS-CoV-2) are infectious, but the extent of transmission based on symptom status requires further study.

**Purpose:** This living review aims to critically appraise available data about secondary attack rates from people with asymptomatic, pre-symptomatic and symptomatic SARS-CoV-2 infection.

**Data sources:** Medline, EMBASE, China Academic Journals full-text database (CNKI), and pre-print servers were searched from 30 December 2019 to 3 July 2020 using relevant MESH terms.

**Study selection:** Studies that report on contact tracing of index cases with SARS-CoV-2 infection in either English or Chinese were included.

**Data extraction:** Two authors independently extracted data and assessed study quality and risk of bias. We calculated the secondary attack rate as the number of contacts with SARS-CoV-2, divided by the number of contacts tested.

**Data synthesis:** Of 927 studies identified, 80 were included. Summary secondary attack rate estimates were 1% (95% CI: 0%-2%) with a prediction interval of 0-10% for asymptomatic index cases in 10 studies, 7% (95% CI: 3%-11%) with a prediction interval of 1-40% for pre-symptomatic cases in 11 studies and 6% (95% CI: 5%-8%) with a prediction interval of 5-38% for symptomatic index cases in 40 studies. The highest secondary attack rates were found in contacts who lived in the same household as the index case. Other activities associated with transmission were group activities such as sharing meals or playing board games with the index case, regardless of the disease status of the index case.

**Limitations:** We excluded some studies because the index case or number of contacts were unclear.

**Conclusion:** Asymptomatic patients can transmit SARS-CoV-2 to others, but our findings indicate that such individuals are responsible for fewer secondary infections than people with symptoms.

**Systematic review registration:** PROSPERO CRD42020188168

**Funding:** No funding was received

## Introduction

Severe Acute Respiratory Syndrome Coronavirus 2 (SARS-CoV-2) demonstrates efficient transmission in populations without effective public health interventions; basic reproduction numbers (R_0_) values range between 2-3 [1]. While asymptomatic transmission has been described as the “Achilles’ heel” of control efforts during this pandemic, the extent to which transmission of SARS-CoV-2 by people without symptoms drives this pandemic remains uncertain [2]. SARS-CoV-2 infection that is asymptomatic at the time of laboratory testing is widely reported [3]; however, studies that follow infected people over time suggest that many infections are not asymptomatic throughout the entire disease course, and a large proportion of these individuals ultimately develop a diverse range of symptoms [4-7]. For instance, Sugano et al. reported a detailed cluster outbreak in music clubs in Japan, where asymptomatic cases reported also included pre-symptomatic cases [8]. A living systematic review of studies published up to 10 June 2020 estimated that 20% (95% CI 17 to 25%) of people who become infected with SARS-CoV-2 remain asymptomatic throughout infection [7].

One of the barriers to understanding the role of asymptomatic transmission is the lack of consistency in case definitions [9]. While symptom severity exists on a spectrum, individuals infected with SARS-CoV-2 can be miscategorized as asymptomatic, when they have milder or atypical symptoms leading to overestimation of the proportion without symptoms [3, 10]. For instance, in a detailed study of SARS-CoV-2 infections in Iceland where individuals deemed at high risk for COVID-19 including those with a consistent syndrome were screened in a targeted manner, and other individuals were tested via a population screening mechanism, more than a third in the second group reported symptoms potentially consistent with COVID-19 [3]. However, it is increasingly becoming clear that some individuals experience more diverse symptoms, including taste and smell disturbance or myalgia, either for the entire course of illness or preceding respiratory symptoms. These symptoms can be so mild and insidious that they do not limit patients’ daily activities [4, 11]. The situation is further complicated by subjective patient perception and differences between studies in the elicitation and reporting of symptoms.

There are reports describing asymptomatic individuals with SARS-CoV-2 who are infectious [12] and who have infected one or more contacts [13], but the extent and significance of asymptomatic transmission requires further understanding. The aim of this review is to summarize the available evidence about secondary attack rates (defined as the probability that an infected individual will transmit the disease to a susceptible individual) amongst the contacts of individuals with SARS-CoV-2 with different symptom status to provide information about how contagious they are, and their role in driving the pandemic.

## Methods

Systematic review was registered in PROSPERO on 8 June 2020 (CRD42020188168) and will be updated 4-6 months according to the availability of new evidence as a living systematic review [14]. The larger review aims to answer transmission dynamics of SARS-CoV-2. The analysis in this report addresses one of the review questions; to identify secondary attack rate based on symptom status.

### Definitions

We defined “asymptomatic” as an individual with laboratory-confirmed SARS-CoV-2 infection but without symptoms throughout their entire course of infection, or after 14 days of follow up; “paucisymptomatic” as an individual with laboratory-confirmed SARS-CoV-2 infection with mild symptoms, and “pre-symptomatic” as an individual who reports no symptoms at the time of the initial positive test result, but who subsequently develops symptoms attributable to COVID-19. We used these definitions to categorize the index cases. Secondary attack rate was defined as the number of new SARS-CoV-2 infection cases among susceptible contacts of primary cases divided by the total number of susceptible contacts.

### Search Strategy

We retrieved articles about transmission of SARS-CoV-2 infection through systematic searches of eight databases: Medline, EMBASE, Europe PMC, Web of Science, SCOPUS, Chinese database (CNKI), and preprint servers (MedRxiv, BioRxiv) using relevant Medical Subject Headings (MeSH) terms (Supplementary material). The initial search was completed from 30 December 2019 to 21 May 2020, searches were repeated on 8 June 2020 and 3 July 2020, owing to the rapidly increasing numbers of studies.

### Study Selection

Studies were eligible if they met the inclusion criteria: (1) report on Coronavirus disease 2019 (COVID-19) or SARS-CoV-2 infection and (2) report an outbreak investigation or contact tracing study. Exclusion criteria were: (1) review articles; (2) observational studies providing only the proportion of individuals infected; (3) studies that do not indicate the number of contacts or secondary infections; and (4) reports in media sources. We also manually screened the references of the included original studies and reviews to identify additional eligible studies.

### Data Extraction

Two authors (XQ and AIN) independently reviewed reports by title and abstract for relevance, with at least 20% of all reports being screened in duplicate to ensure consistency. Two authors then independently read the full text report of all studies not excluded by title and abstract, to consider eligibility for inclusion. Any disagreements regarding study inclusion were resolved through discussion with a third author (MC). Data were extracted onto a standardized form. From each study, the following variables were extracted: the name of the first author, year of publication, country, sample size, details of index cases (categorised as asymptomatic, pre-symptomatic and symptomatic); event details such as environment, transmission details; number of contacts, number of secondary cases. If these data were not reported, we contacted authors to request them and checked with the authors about all symptoms that they sought.

### Risk of bias in included studies

Two authors (XQ and AIN) independently assessed completeness of reporting and risks of bias, using an adapted version of the Joanna Briggs Institute Critical Appraisal Checklist for Case Series (Supplementary material). Any disagreements were resolved through discussion with a third author (MC).

### Data synthesis and statistical analysis

The studies are summarized in text and table form, descriptive statistics were completed for key outcome measures. Secondary attack rates were computed from raw data in each study, dividing the number of infected contacts of primary cases by the total number of susceptible exposed contacts. A pooled analysis was carried out to generate summary estimates for the secondary attack rate in each subgroup analyzed (asymptomatic, pre-symptomatic and symptomatic index cases), in the framework of a random effect model. The Freeman – Tukey double-arcsine variance-stabilizing transformation was used to combine data, due to its advantage over log and logit transformations which did not allow to compute the proportion in the presence of zero event counts [15]. Secondary attack rates are presented as a proportion along with 95% CIs in forest plots. Heterogeneity between study estimates was gauged by means of the Cochran’s Q and I^2^ statistic: an I^2^ value above 75% indicates high heterogeneity [16]. Moreover, a 95% prediction interval is displayed in the forest plots, which is an index of dispersion, providing information on how widely the true effect size varies. It can also provide the range of values in which a future observation will fall [17]. Analyses were performed though the software MetaXL version 5.3 (Ersatz, EpiGear International, Sunrise Beach, Australia) [18].

## Results

The systematic search identified 927 potentially relevant articles and 789 records were screened after removal of duplicates. Of 187 articles retrieved for full-text review and assessed for eligibility, 80 studies were included in the systematic review, and among those we identified 69 studies that indicated the symptom status of index case(s). In this analysis, we excluded 11 studies that reported asymptomatic and symptomatic index cases together or no symptom status of the index case was available. We re-classified three studies from asymptomatic to pre-symptomatic as the index cases developed symptoms later during the disease course after reviewing the details and contacting the authors [19-21]. The number of selected papers at each step of the screening and eligibility are reported in the flow diagram (Figure 1).

**Figure 1.**
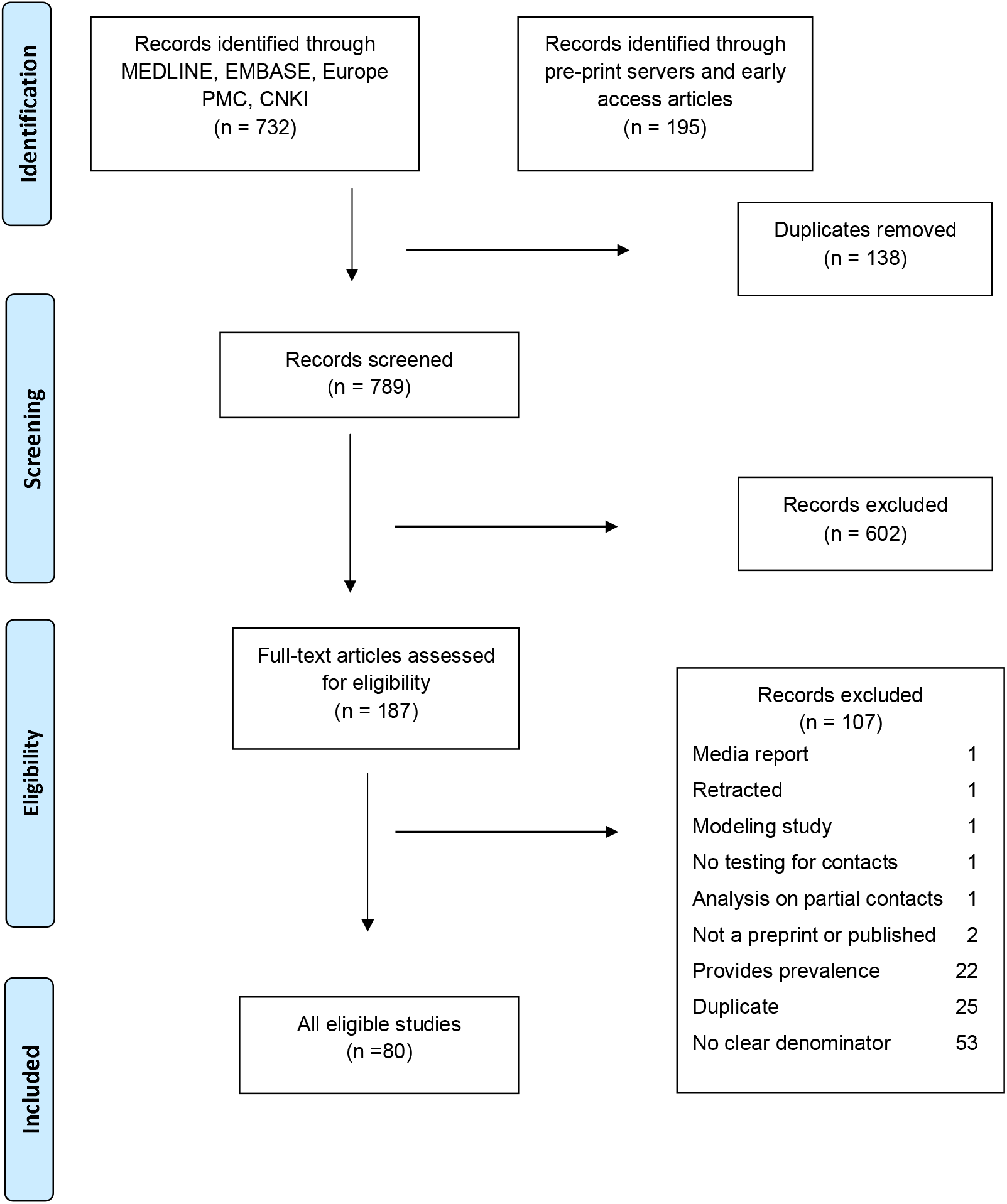
Flowchart describing inclusion and exclusion of studies at each stage of the review.

### Summary of secondary attack rates of asymptomatic index cases

Ten studies were included in the quantitative analysis (Table 1) [6, 13, 22-29]. Summary secondary attach rate estimate was 1% (95% CI: 0%-2%) with a prediction interval ranged of 0-10% (Figure 2). All except one tested all close contacts for SARS-CoV-2, regardless of symptoms [26]. Cheng et al. only tested symptomatic cases, but they also tested high risk populations regardless of symptoms including the household and hospital contacts [26]. Six studies reported on household contacts, two studies included hospital contacts and two studies included non-household close contacts.

**Table 1:**
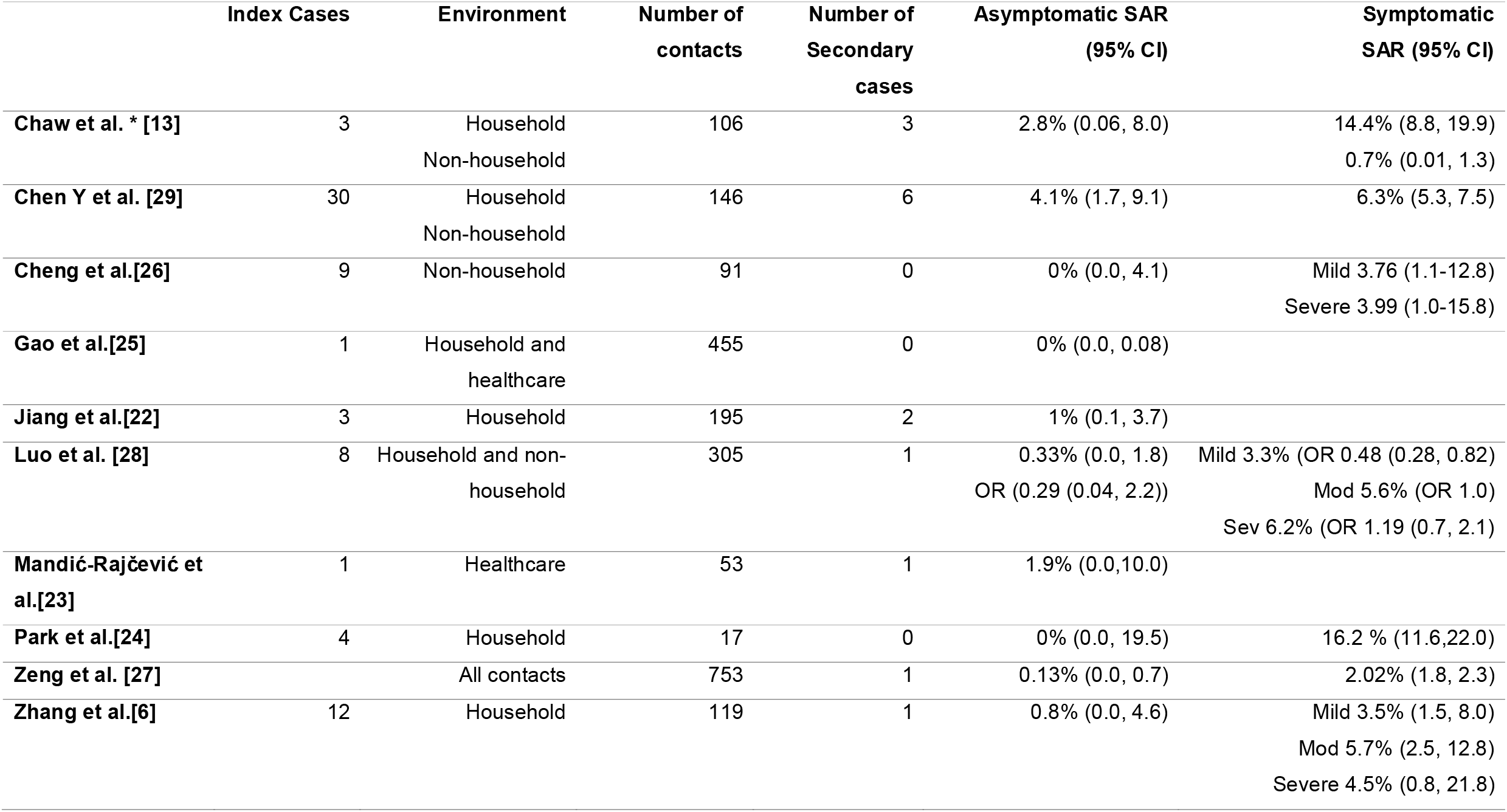
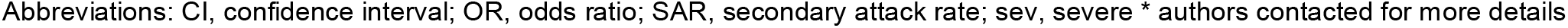
Transmission from truly asymptomatic index cases

**Figure 2:**
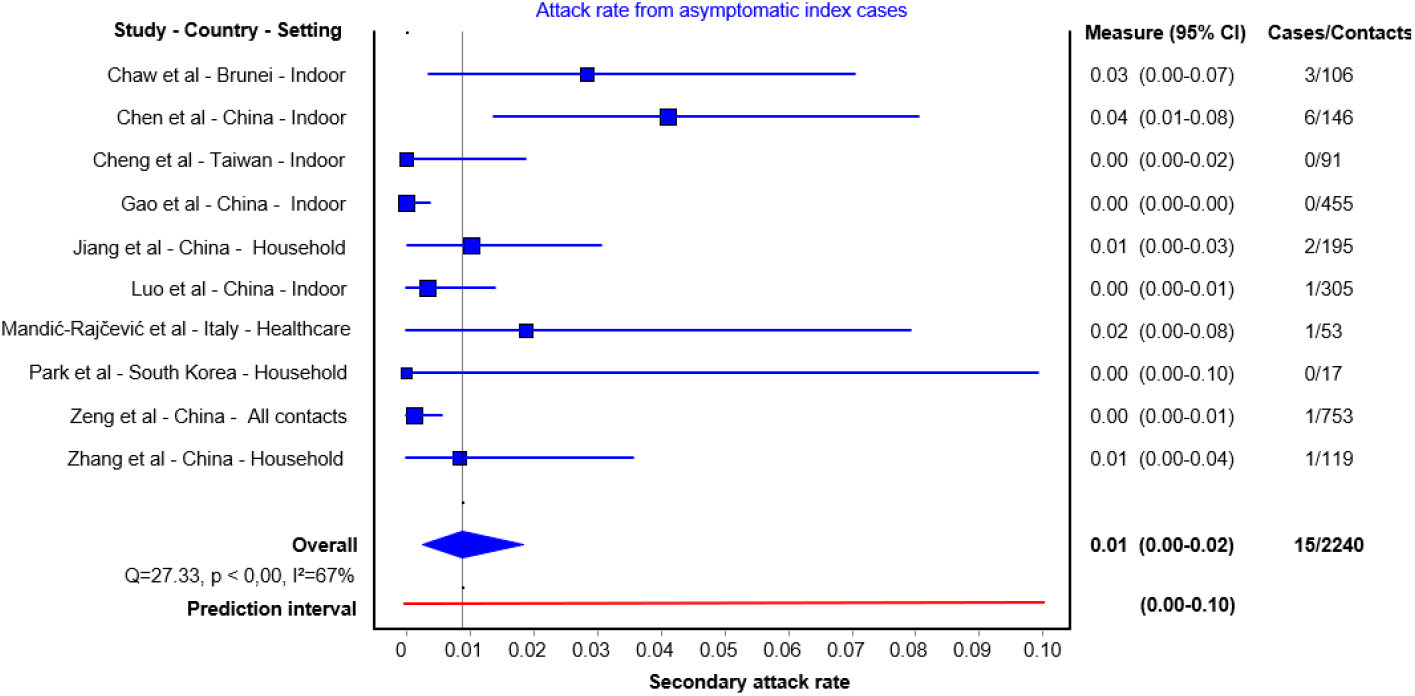
Secondary attack rates from asymptomatic index cases to their contacts. For each study the secondary attack rate is reported with its 95% CI. A prediction interval at the bottom of the forest is depicted.

Three studies identified no secondary cases after following up 17, 91 and 455 close contacts of asymptomatic index cases (asymptomatic secondary attack rate of 0%) [24-26]. Of those, two studies demonstrated higher symptomatic secondary attack rates; Cheng et al. demonstrated that mild cases had a secondary attack rate of 3.8% (95% CI 1.1, 12.8%) and severe cases had 4% (95% CI 1.0, 15.8%) secondary attack rate [26], while Park et al. showing household symptomatic secondary attack rate of 16.2% (95% CI 11.6, 22.0%) [24]. In another study, 305 contacts of 8 asymptomatic cases were followed up, identifying one secondary case (secondary attack rate 0.3% (95% CI 0.0, 1.8%) [28]. In the same study, attack rates from index cases with mild, moderate and severe diseases were 3.3%, 5.6% and 6.2%, respectively. Zhang et al. followed up 119 close contacts of 12 asymptomatic index cases and identified one secondary case, an asymptomatic secondary attack rate of 0.8% (95% CI 0.0, 4.6). In the same study, the secondary attack rate was 3.5% (95% CI 1.5-8.0) for those with mild, 5.7% (95% CI 2.5, 12.8%) for those with moderate, and 4.5% (95% CI 0.8, 21.8%) for those with severe symptoms [6]. In this study, close contacts that lived with an index case had 12 times the risk of infection as those who did not live with the index case (RR 12.5 - 95% CI 1.6, 100.8) and those who had frequent contact with an index case-patient, and those who had more than 5 contacts had 29 times the risk of infection as those with fewer contacts (RR 29.0 - 95% CI 3.6, 232.3). Two studies indicated an asymptomatic secondary attack rate of 1% and 1.9% [22, 23]. Chaw et al. reported asymptomatic and pre-symptomatic contacts together. The authors clarified that 3 asymptomatic index cases and their 106 close contacts were followed up, leading to 3 secondary cases, a secondary attack rate of 2.8% (95% CI 0.06, 8.0%). In this study, the overall secondary attack rate was 10.6% in the household setting, which was higher for symptomatic cases (14.4%, 95% CI 8 · 8, 19 ·9%) than that of asymptomatic cases and for non-household contacts 0.7 (95% CI 0.1, 1,3) [13]. Zeng et al. conducted the largest contact tracing study, following up 753 close contacts of asymptomatic index cases and identified one secondary case, an asymptomatic secondary attack rate of 0.13% (95% CI 0.0, 0.7%) [27].

### Summary of pre-symptomatic secondary attack rates

Sixteen papers reported either outbreak investigations or contact tracing studies reporting transmission from an index case during the pre-symptomatic period [13, 19, 21, 26, 30-40] (Table 2). Of those, eleven studies were included in the quantitative analysis. The summary SAR estimate was 7% (95% CI: 3%-11%) with a prediction interval of 1 to 40% (Figure 3). These studies followed up 22 to 585 close contacts whose initial exposure occurred before symptom onset of the index case. Even in studies that followed up large numbers of people, including community contacts, the majority of secondary cases identified were from the same household or among friend gatherings. In these studies, having meals together, or playing cards with the index case were exposure activities associated with transmission. The remaining one study reported an outbreak in a restaurant [40] and four studies exclusively reported family cluster outbreaks [30, 32, 33, 39]; these investigations did not test contacts outside the household, and it is challenging to truly differentiate transmission during the pre-symptomatic period from symptomatic transmission in the household setting (Supplementary Figure 1).

**Table 2:** Transmission during pre-symptomatic period

| **Index**<br>**Cases** | **Environment** | **Number of**<br>**contacts** | **Number of**<br>**Secondary**<br>**cases** | **Pre-symptomatic**<br>**SAR (95% CI)** | **Secondary cases** |
| --- | --- | --- | --- | --- | --- |

**Table 2: Transmission during pre-symptomatic period** Transmission dynamics and secondary attack rates
| Contract-tracing |  |  |  |  |  |  |
| --- | --- | --- | --- | --- | --- | --- |
| Chaw et al.* [13] | 7 | Household and non-household | 585 | 15 | 2.56% (1.4, 4.2) |  |
| Cheng et al.[26] | NR | Household and non-household | 299 | 2 | 0.7% (0.1, 2.4) |  |
| Hong L et al.[31] | 41 | Household and non-household | 197 | 24 | 12.2% (8.0, 17.6) | Friends, family, card playing partners |
| Huang et al.[19] | 1 | Friends | 22 | 7 | 31.8% (13.0, 54.9) | Shared meal with index |
| Pang et al. [34] | 1 | Household and non-household | 103 | 6 | 5.8% (2.2, 12.2) | Living together or sharing meal |
| Park et al. [24] | 4 | Household | 11 | 0 | 0% (0.0, 2.8) |  |
| Pung R et al. [38] | 2 | Religious gathering | 142 | 3 | 2.1% (0.5, 6.5) |  |
| Qian et al. [35] | 1 | Household and non-household | 137 | 10 | 7.3% (3.6, 13.0) | Living together or sharing meal |
| Yang et al. [36] | 2 | Household and non-household | 123 | 6 | 4.9% (1.8, 10.3) | All secondary cases lived together |
| Ye et al. [21] | 1 | Family | 44 | 4 | 9.1% (2.5, 21.4) | Extended family |
| Zhao et al [37] | 1 | Friends | 15 | 4 | 26.7% (7.8, 55.1) | Meal and Mahjong game gathering |
| Outbreak investigation |  |  |  |  |  |  |
| Chen M et al.[30] | 1 | Household | 3 | 2 | 66.7% (9.4, 99.2) | Family cluster outbreak |
| Li P et al.[32] | 1 | Household | 5 | 4 | 80% (28.4, 99.5) | Family cluster outbreak |
| Lu J et al [40] | 1 | Restaurant | 82 | 9 | 11% (5.9, 19.6) |  |
| Jiang Y et al [39] | 1 | Household | 7 | 3 | 42.9% (11.8, 79.8) | Family cluster outbreak |
| Qian G et al. [33] | 2 | Household | 4 | 3 | 75% (10.4, 99.4) | Family cluster outbreak |

**Figure 3:**
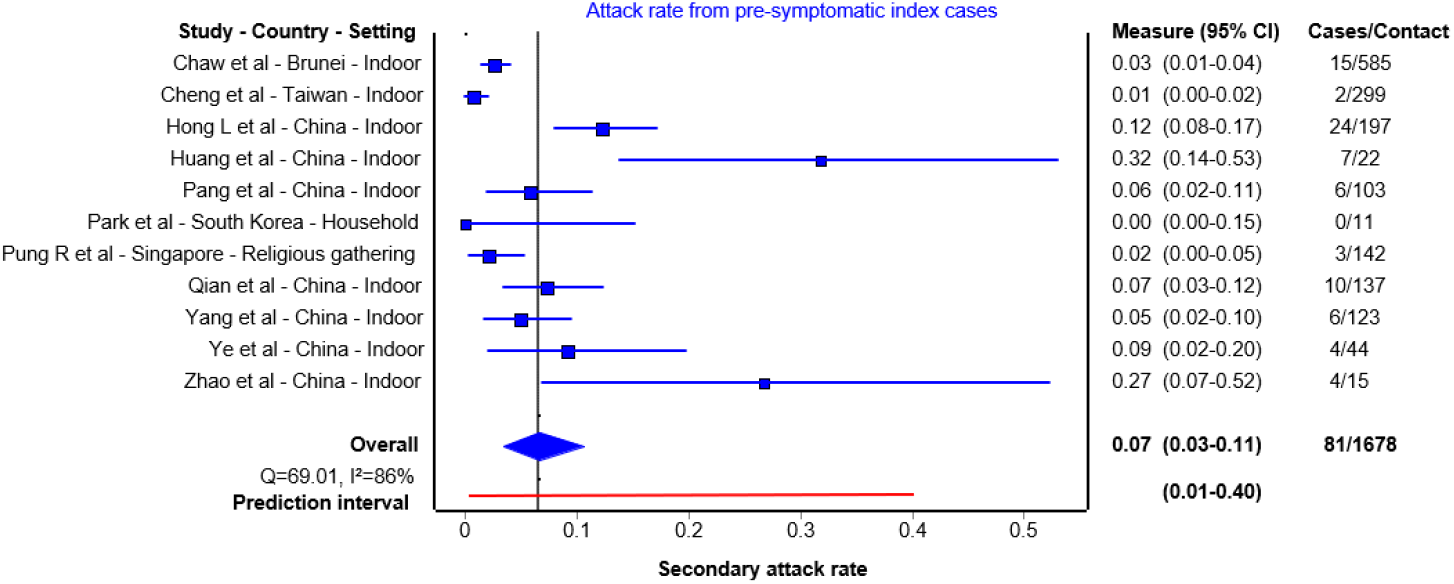
Secondary attack rates from pre-symptomatic index cases to their contacts. For each study the secondary attack rate is reported with its 95% CI. A prediction interval at the bottom of the forest is depicted.

### Summary of symptomatic secondary attack rates

Forty-six papers reported either outbreak investigations or contact tracing studies reporting transmission from symptomatic index case(s). Of those, 40 reported contact tracing studies reported secondary attack rates ranging from 0% to 38.89% [13, 24, 27, 28, 38, 41-75] and 6 reported outbreak investigation [76-80] (Supplementary Table 1). 40 contact tracing studies with 44 observations were included in the quantitative analysis (Figure 4). The summary estimate of SAR from symptomatic index subjects was 6% (95% CI: 5%-8%) with a prediction interval of 5- 38%. Of those, 9 studies reported less than 1% secondary attack rates, 2 of those were in healthcare setting, 2 included outdoor interaction, 4 included non-household contacts. Higher frequency of contacts and household contacts were reported to be higher risk than non-household contacts.

**Figure 4:**
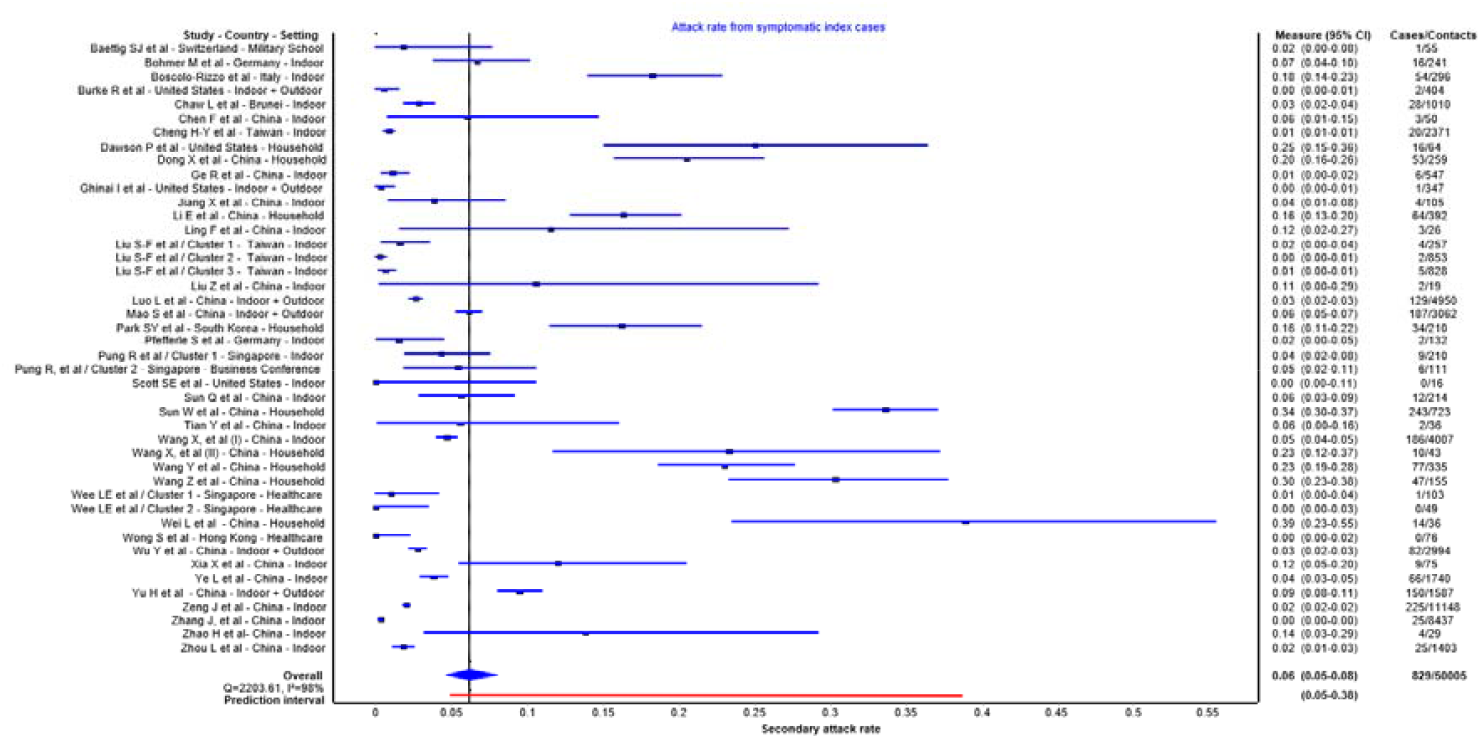
Secondary attack rates from symptomatic index cases to their contacts. For each study the secondary attack rate is reported with its 95% CI. A prediction interval at the bottom of the forest is depicted.

### Quality assessment

All papers included a clear definition of symptomatic and asymptomatic cases, number of secondary cases and number of contacts. The majority of studies identified index cases with a clear diagnosis, had an acceptable case definition and sufficiently followed up close contacts (for a minimum of 14 days). However, in some studies the definition of close contact and setting of transmission was not provided. In addition, it was unclear in four reports whether all potential close contacts were included, therefore, the direction of bias is uncertain. We summarized the quality assessment in Supplementary Table 2.

## Discussion

This systematic review provides comprehensive data on secondary attack rates based on symptom status of the index case(s). While asymptomatic patients can transmit the virus to others [81], the findings from ten studies in this review found summary secondary attack rates of 1% with a prediction interval of 0-10% for asymptomatic index cases compared with secondary attack rates of 6% with a prediction interval of 5-38% in symptomatic cases and 7% with a prediction interval of 1-40% in pre-symptomatic case. These findings suggest that individuals who are asymptomatic throughout the disease course are responsible for fewer secondary infections than symptomatic and pre-symptomatic cases. Most transmission events were associated with living with the index case or group activities such as sharing meals and playing board games.

Given the importance of transmission heterogeneity in propagating the pandemic, it is important that we learn about the various factors that contribute to transmission. According to modelling and contact tracing studies, around 80% of secondary infections can be linked to 20% of cases which distinguishes SARS-CoV-2 from seasonal influenza, although a similar pattern was also observed in SARS-CoV and MERS-CoV [82-84]. While there are multiple factors (environmental factors, contact patterns and socioeconomic inequalities) that contribute to this heterogeneity, some evidence starting to emerge about the influence of individual’s infectiousness on transmission dynamics. In this systematic review, we found that index cases with symptoms had a higher secondary attack rate compared with truly asymptomatic index cases. While there is a need to better understand this difference, it may be due to shorter duration of infectiousness. In a living systematic review including of studies published up to 6 June 2020, we found that cases with asymptomatic people had a shorter duration of RNA shedding than symptomatic individuals [85]. Asymptomatic patients may therefore be contagious but for a shorter duration than symptomatic people; this might contribute to lower transmission to their contacts. However, we do not yet know the relative importance of behavioral factors by the host versus environmental factors in determining transmission risk. It is not known whether the size of the cluster of secondary infections would be different according to index case symptom status in a high-risk environment with no mitigation measures in place.

Modelling studies suggest that it is not possible to have widespread infection without substantial pre-symptomatic transmission. Viral load dynamics of SARS-CoV-2 derived from confirmed cases suggest that peak viral loads are detected at the start of symptom onset up to day 5 of illness, indicating highest infectiousness occurs just before or within the first few days after symptom onset [85]. So far, several contact tracing studies emphasize that the highest risk of transmission occurs during the prodromal phase or early in the disease course [64, 86]. For instance, in a prospective contact tracing study of 100 confirmed cases of COVID-19 and 2761 close contacts, no secondary cases were identified when the exposure occurred more than 5 days after the symptom onset [26]. Our findings therefore have important implications from a public health perspective. In settings such as nursing homes, homeless shelters, prisons, cruise ships and meat-packing plants in which many people spend prolong period of time together in the same environment including sleeping, dining and sharing common facilities, and where several outbreaks have been documented, pre-symptomatic transmission may contribute substantially to transmission [87, 88]. In these settings, when infection develops, most patients are already inside the facility with high viral loads that increase the risk of onward transmission. This highlights the importance of mitigation measures and surveillance in these settings to identify those patients early in the disease course to prevent onward transmission inside the facility.

This systematic review has several strengths. Firstly, this is a living systematic review examining the transmission of SARS-CoV-2 through contact tracing and outbreak investigation studies. Secondly, we only included studies with clear case definitions, which indicated the number of contacts and secondary cases. We excluded studies in which the index case was unclear, or the numbers of contacts were not provided. The estimates from individual studies are also subject to limitations, such as imprecision resulting from small study size, and multiple sources of bias in the estimation of the true secondary attack rate, which are detailed in this paper [89]. Moreover, while the number of index cases could influence the confidence interval estimation for secondary attack rate due to heterogeneity among index cases, we have constructed a prediction interval to yield conservative confidence interval estimates.

We identified two other systematic reviews that investigated asymptomatic transmission, with different research questions, which results in different search terms and studies retrieved. One living systematic review, which included studies published up to 10 June 2020, identified five studies that directly compared secondary attack rates between asymptomatic and symptomatic index cases; all were included in our review [7]. This study only included studies that provided data to allow relative risks to be estimated. The summary risk ratios for asymptomatic versus symptomatic (0.35, 95% CI 0.10, 1.27) and pre-symptomatic versus symptomatic (0.63, 95% CI 0.18, 2.26) are consistent with our findings. The second review estimated only household secondary attack rates and included studies published up to 29 July 2020 [90]. Of three studies that included asymptomatic index cases, two were included in our review. We excluded one of the studies because the number of contacts of asymptomatic index cases was not specified; we have not yet received details of the study after contacting the authors. Advantages of our review over these two studies are inclusion of studies published in Chinese, search terms that aimed to capture studies specifically estimating secondary attack rates.

In summary, whilst asymptomatic transmission is a major concern for SARS-CoV-2 community spread, secondary attack rates from those who remain asymptomatic throughout their course of infection are low suggesting limited infectiousness. Although it is difficult to estimate the proportion of pre-symptomatic transmission, these patients are likely to be highly infectious just before and around the time of symptom onset and appear to transmit efficiently, particularly within households. Given these results, in the context of limited resources, approaches should be targeted predominantly on identifying and immediately isolating patients with prodromal or mild symptoms and their contacts, which may avert a significant number of community transmission clusters [91]. Future clinical studies should incorporate clear definitions and assess a broad range of symptoms associated with COVID-19, include longitudinal follow up of patients, and calculate secondary attack rates for a wider range of settings and populations [9].

## Data Availability

Not applicable

## Acknowledgements

We would like to thank the authors of Chau et al. (Dr Tan Le Van), Mandić-Rajčević et al. (Dr Stefan Mandić-Rajčević), Chaw et al. (Dr Liling Chaw) for providing further details about asymptomatic cases in their reports also Prof Stephen Gillespie for his comments on the first draft of this manuscript. We would like to acknowledge Dr. Shuang Jin for searching and downloading Chinese database for this review.

## Contribution statement

X Qiu: investigation, data curation, writing – original draft; A Nergiz: investigation, data curation, writing – review and editing; A. E. Maraolo: methodology, formal analysis, writing – original draft; I. Bogoch and N. Low: interpretation, writing – review and editing; M. Cevik: conceptualisation, methodology, investigation, writing – original draft, supervision.

