## Supplementary material for "The role of asymptomatic and pre-symptomatic infection in SARS-CoV-2 transmission – a living systematic review"

**Differences in secondary attack rates based on symptom status of index case(s)– a living systematic review**

**Author of correspondence:**

Name: Dr Muge Cevik

Telephone number: +447732800814

**Supplementary Figure 1: Secondary attack rates from pre-symptomatic index cases to their contacts in outbreak investigations (not contact tracing studies).**

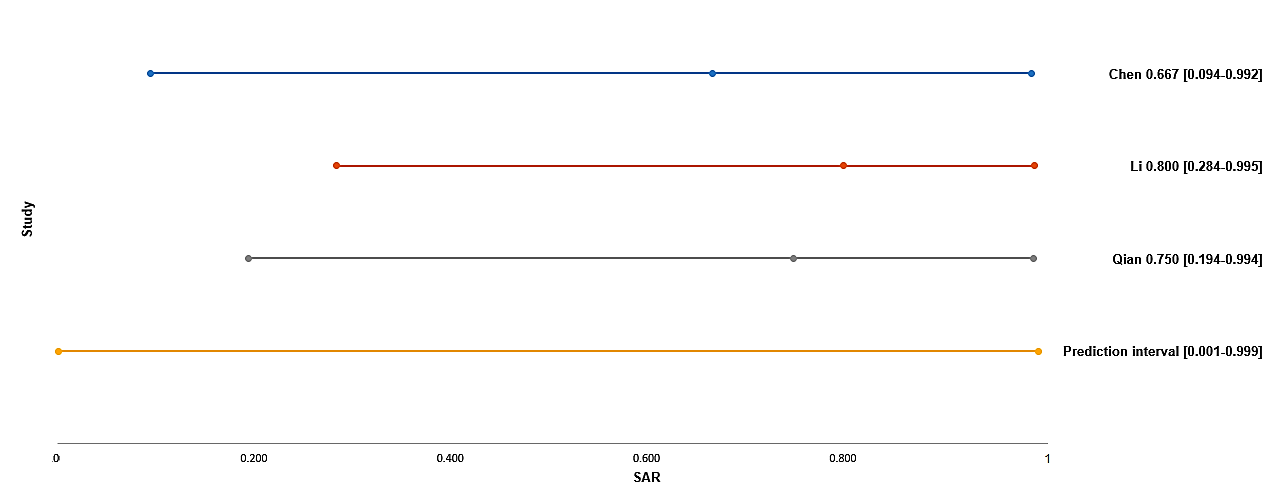

For each study the secondary attack rate is reported with its 95% CI.

A prediction interval at the bottom of the forest is depicted.

**Supplementary Table 1: Transmission from symptomatic index cases**

| Study ID | Study | Environment | Number of index cases | Number of contacts | Number of secondary cases | Secondary attack rate | Risk factors associated with high SAR |
| --- | --- | --- | --- | --- | --- | --- | --- |
| Q12 | Baettig SJ, et al | Military school | 1 | 55 | 1 | 1.82% |  |
| Q20 | Bohmer M, et al | Household and non-household | 1 | 241 | 16 | 6.64% |  |
| A23 | Boscolo-Rizzo P, et al | Household | 179 | 296 | 54 | 18.24% | Shared meal |
| A2 | Burke R, et al | Indoor + Outdoor | 9 | 404 | 2 | 0.50% |  |
| A4 | Chaw L, et al | Household and non-household | 19 | 1010 | 28 | 2.77% |  |
| Q43 | Chen F, et al | Household and non-household | 1 | 50 | 3 | 6.00% | Household, bus |
| A1 | Cheng H-Y, et al | Household and non-household | 100 | 2371 | 20 | 0.84% | The attack rates were higher among those aged 40 and older; Severe condition in index case |
| A24 | Dawson P, et al | Household | 24 | 64 | 16 | 25.00% | Shared meal |
| Q30 | Dong X, et al | Household | 26 | 259 | 53 | 20.46% | Shared meal |
| A3 | Ge R, et al | Household and non-household | 1 | 547 | 6 | 1.10% |  |
| Q18 | Ghinai I, et al | Indoor + Outdoor | 1 | 347 | 1 | 0.29% | Living together |
| Q21 | Jiang X, et al | Household and non-household | 8 | 105 | 4 | 3.81% |  |
| A6 | Li W, et al | household | 105 | 392 | 64 | 16.33% | Being adult compared to children, being spouse of index case compared to other adults, shared meal |
| Q32 | Ling F, et al | Household and healthcare | 1 | 26 | 3 | 11.54% | Household, healthcare |
| A13 | Liu S-F, et al - Cluster 1 | Household and non-household | 1 | 257 | 4 | 1.56% |  |
| A13 | Liu S-F, et al - Cluster 2 | Household and non-household | 1 | 853 | 2 | 0.23% |  |
| A13 | Liu S-F, et al - Cluster 3 | Household and non-household | 1 | 828 | 5 | 0.60% |  |
| Q33 | Liu Z, et al | Household and non-household | 1 | 19 | 2 | 10.53% | Living together; sharing meals |
| A28 | Luo L, et al | Indoor + Outdoor | 347 | 4950 | 129 | 2.61% |  |
| A21 | Mao S, et al | Indoor + Outdoor | 75 | 3062 | 187 | 6.11% |  |
| A8 | Park SY, et al | Household | 94 | 210 | 34 | 16.19% | Living together |
| Q25 | Pfefferle S, et al | Household and non-household | 1 | 132 | 2 | 1.52% |  |
| A11 | Pung R, et al - Cluster 1 | Household and non-household | 5 | 210 | 9 | 4.29% |  |
| A11 | Pung R, et al - Cluster 2 | Business Conference | 1 | 111 | 6 | 5.41% | Shared meals |
| Q5 | Scott SE, et al | Household and non-household | 1 | 16 | 0 | 0.00% | Index case has really mild symptoms – low risk |
| Q35 | Sun Q, et al | Household and non-household | 1 | 214 | 12 | 5.61% |  |
| Q16 | Sun W, et al | Household | 148 | 723 | 243 | 33.61% | Shared meal |
| Q40 | Tian Y, et al | Household and non-household | 1 | 36 | 2 | 5.56% | Shared meal |
| Q29 | Wang X, et al | Household | 25 | 43 | 10 | 23.26% | Shared meal |
| Q2 | Wang X, et al | Household and non-household | 416 | 4007 | 186 | 4.64% | Household, healthcare workers |
| A25 | Wang Y, et al | Household | 41 | 335 | 77 | 22.99% | Shared meal; having frequent daily contact with index case, index case with diarrhoea, using less face mask |
| Q19 | Wang Z, et al | Household | 78 | 155 | 47 | 30.32% | Shared meal |
| A15 | Wee LE, et al - Cluster 1 | Healthcare | 1 | 103 | 1 | 0.97% |  |
| A15 | Wee LE, et al - Cluster 2 | Healthcare | 2 | 49 | 0 | 0.00% |  |
| Q7 | Wei L, et al | Household | 60 | 36 | 14 | 38.89% | Immediate family; living together, duration of exposure |
| Q23 | Wong S, et al | Healthcare | 1 | 76 | 0 | 0.00% | PPE protective measures in place |
| A26 | Wu Y, et al | Indoor + Outdoor | 144 | 2994 | 82 | 2.74% |  |
| A17 | Xia X, et al | Household and non-household | 1 | 75 | 9 | 12.00% | Having dinner |
| Q9 | Ye L, et al | Household and non-household | 1 | 1740 | 66 | 3.79% | Bus |
| A30 | Yu H, et al | Indoor + Outdoor | 560 | 1587 | 150 | 9.45% |  |
| Q39 | Zeng J, et al | Household and non-household | 313 | 11148 | 225 | 2.02% |  |
| Q11 | Zhang J, et al | Household and non-household | 1 | 8437 | 25 | 0.30% | Family members and relatives |
| Q10 | Zhao H, et al | Friends | 1 | 29 | 4 | 13.79% | Gathering + board game + meals |
| Q34 | Zhou L, et al | Household and non-household | 116 | 1403 | 25 | 1.78% | Higher frequency of contacts (SAR = 22/129 = 17.05%); home (24/165 = 14.55%); living together (24/100=24%); |
| Mixed index case status | | | | | | | |
| A19 | Bi Q, et al | Household and non-household | 391 | 1286 | 98 | 7.62% | Living/travel together; shared meals |
| Q31 | Chen Y, et al | Household and non-household | 187 | 2147 | 132 | 6.15% |  |
| Q24 | Jia H, et al | Household and non-household | 73 | 1159 | 24 | 2.07% | Old people under care, family members, medical staff |
| Q15 | Jing Q, et al | Household and non-household | 349 | 1978 | 360 | 18.20% | Household |
| A20 | Kwok KO, et al | Indoor + Outdoor | 53 | 206 | 24 | 12.75% |  |
| A29 | Son H, et al | Household | 108 | 196 | 16 | 8.16% | Shared meal |
| Q1 | Wu J, et al | Household | 35 | 148 | 48 | 32.43% | Shared meal |
| Unclear index case status | | | | | | | |
| Q6 | Jing Q, et al | Household and non-household | 212 | 2075 | 137 | 6.60% | Older age, household |
| Q38 | Jing R, et al | Household and non-household | 3 | 777 | 5 | 0.64% | Workplace and household |
| Q4 | Korea CDC | Household and non-household | 30 | 2370 | 13 | 0.55% |  |
| Q37 | Zhang R, et al | Indoor + Outdoor | / | 2784 | 67 | 2.41% | Living together, staying in the same space or having meals together. |
| Outbreak Investigation | | | | | | | |
| A22 | Chen D, et al | Household | 1 | 5 | 3 | 60% | Living together |
| Q14 | Ghinai I, et al | Birthday gathering | 1 | 9 | 3 | 33.33% | Birthday party, shared meal |
| Q17 | Huang R, et al | Household | 1 | 4 | 3 | 75% | Living together and dinner gathering |
| Q13 | Li C, et al | Household | 1 | 4 | 2 | 50% | Living together |
| A7 | Song R, et al | Household | 4 | 20 | 18 | 90% | Living together |
| A12 | Tong Z-D, et al | household | 2 | 4 | 3 | 75% | Living together |
| A18 | Xu Y, et al | Household | 2 | 4 | 4 | 100% | Living together |

**Abbreviation: SAR – secondary attack rate.**

**Supplementary Table 2: Quality assessment of studies**

| **Study ID** | **Author** | **1. Was the research question or objective in this paper clearly stated?** | **2. Was the study population clearly specified and defined?** | **3. Was the index case(s) well identified with clear diagnose or demographic information?** | **4. Was an acceptable definition of (household, hospital) close contacts used in the study?** | **5. Was it including/representing all potential close contacts of the index case(s) based on study question?** | **6. Was the settings/environment of the infections clear?** | **7. Was an acceptable case definition used in the study (numerator for the research question)?** | **8. Was the length of follow-up appropriate to get final outcome of all contacts?** |
| --- | --- | --- | --- | --- | --- | --- | --- | --- | --- |
| Q12 | Baettig, et al | Y | Y | Y | Y | Y | Y | Y | Y |
| A19 | Bi Q, et al | Y | Y | Y | Y | Y | Y | Y | Y |
| Q20 | Bohmer, et al | Y | U | Y | Y | Y | U | Y | Y |
| A23 | Boscolo‑Rizzo, et al | Y | Y | Y | Y | Y | Y | Y | U |
| A2 | Burke, et al | Y | Y | Y | Y | Y | Y | Y | Y |
| A2* | Burke, et al | Y | Y | U | Y | Y | Y | Y | Y |
| A4 | Chaw L, et al | Y | Y | Y | Y | Y | Y | Y | Y |
| A22 | Chen D, et al | Y | Y | U | Y | Y | Y | Y | Y |
| Q43 | Chen F, et al | Y | Y | Y | U | Y | Y | Y | Y |
| Q22 | Chen M, et al | Y | Y | Y | U | Y | Y | Y | Y |
| Q31 | Chen Y, et al | Y | Y | Y | Y | Y | Y | Y | Y |
| Q31* | Ding, et al | Y | Y | Y | U | Y | Y | Y | U |
| Q31* | Yin G, et al | Y | Y | Y | Y | Y | Y | Y | Y |
| A1 | Cheng H-Y, et al | Y | Y | Y | Y | Y | Y | Y | Y |
| A24 | Dawson, et al | Y | Y | U* | Y | Y | Y | Y | U |
| Q26 | Deng L, et al | Y | Y | U | Y | Y | Y | Y | Y |
| Q30 | Dong X, et al | Y | Y | Y | U | U | U | Y | U |
| Q43 | Gao M, et al | Y | Y | Y | Y | Y | Y | Y | U |
| A3 | Ge R, et al | Y | Y | U | Y | Y | U | Y | Y |
| Q14 | Ghinai, et al | Y | U | Y | U | U | Y | Y | Y |
| Q18 | Ghinai, et al | Y | Y | Y | Y | Y | Y | Y | Y |
| Q27 | Hong L, et al | Y | U | Y | Y | U | Y | Y | U |
| Q8 | Huang L, et al | Y | Y | Y | U | U | Y | Y | Y |
| Q17 | Huang R, et al | Y | Y | U | U | U | Y | U | Y |
| Q24 | Jia H, et al | Y | Y | Y | Y | Y | Y | Y | U |
| Q21 | Jiang X, et al | Y | Y | U | Y | Y | U | Y | Y |
| A5 | Jiang Y, et al | Y | U | Y | N | U | Y | Y | Y |
| Q6 | Jing Q, et al | Y | Y | Y | Y | Y | Y | Y | Y |
| Q6* | Ma Y, et al | Y | Y | Y | Y | Y | Y | Y | Y |
| Q15 | Jing Q, et al | Y | Y | Y | Y | Y | Y | Y | Y |
| Q38 | Jing R, et al | Y | Y | Y | Y | Y | Y | Y | Y |
| Q4 | Korea CDC, et al | Y | Y | U | Y | Y | Y | N | Y |
| A20 | Kwok, et al | Y | Y | Y | Y | Y | U | Y | U |
| Q13 | Li C, et al | Y | Y | Y | U | Y | Y | Y | U |
| A14 | Li P, et al | Y | Y | Y | Y | Y | Y | Y | Y |
| A6 | Li W, et al | Y | Y | Y | Y | Y | Y | Y | Y |
| Q32 | Ling F, et al | Y | Y | Y | Y | Y | Y | Y | Y |
| A13 | Liu S-F, et al | Y | Y | Y | U | Y | U | Y | Y |
| A13* | Liu, et al | Y | Y | Y | U | Y | U | Y | Y |
| Q33 | Liu Z, et al | Y | Y | Y | Y | Y | Y | Y | Y |
| A9 | Lu J, et al | Y | Y | Y | U | Y | Y | Y | Y |
| A28 | Luo L, et al | Y | Y | Y | Y | Y | Y | Y | Y |
| Q3 | Mandic-Rajcevic, et al | Y | Y | Y | Y | U | Y | Y | Y |
| A21 | Mao S, et al | Y | Y | Y | U | Y | U | Y | Y |
| Q36 | Pang Q, et al | Y | Y | Y | Y | Y | U | Y | Y |
| A8 | Park, et al | Y | Y | Y | U | Y | Y | Y | Y |
| Q25 | Pfefferle, et al | U | U | Y | Y | U | Y | Y | Y |
| A11 | Pung R, et al | Y | Y | U | Y | Y | Y | Y | Y |
| A10 | Qian G, et al | Y | U | Y | U | Y | Y | Y | Y |
| Q41 | Qian L, et al | Y | Y | Y | Y | Y | Y | Y | Y |
| Q5 | Scott, et al | Y | Y | Y | Y | Y | Y | U | Y |
| A29 | Son Y, et al | Y | Y | U | Y | Y | Y | Y | Y |
| A7 | Song R, et al | Y | Y | Y | U | Y | Y | Y | Y |
| Q35 | Sun Q, et al | Y | Y | Y | Y | Y | U | Y | Y |
| Q16 | Sun W, et al | Y | Y | Y | Y | Y | Y | Y | U |
| Q40 | Tian Y, et al | Y | Y | Y | Y | Y | Y | Y | Y |
| A12 | Tong Z, et al | Y | Y | Y | U | Y | Y | Y | Y |
| Q2 | Wang X, et al | Y | Y | U | Y | Y | Y | Y | Y |
| Q29 | Wang X, et al | Y | Y | Y | U | Y | Y | Y | U |
| A25 | Wang Y, et al | Y | Y | Y | Y | Y | Y | Y | Y |
| Q19 | Wang Z, et al | Y | Y | Y | Y | Y | Y | Y | Y |
| A15 | Wee L, et al | Y | Y | Y | Y | Y | Y | Y | Y |
| Q7 | Wei L, et al | Y | Y | U | U | U | Y | N | U |
| Q23 | Wong S, et al | Y | Y | Y | Y | Y | Y | Y | Y |
| Q1 | Wu J, et al | Y | Y | Y | Y | Y | Y | Y | Y |
| A26 | Wu Y, et al | Y | Y | U | Y | Y | Y | U | Y |
| A17 | Xia X, et al | Y | Y | Y | Y | Y | U | Y | U |
| A18 | Xu Y, et al | Y | Y | U | Y | Y | Y | Y | U |
| Q42 | Yang Y, et al | Y | Y | Y | Y | Y | Y | Y | Y |
| A16 | Ye F, et al | U | Y | Y | U | U | U | Y | U |
| Q9 | Ye L, et al | Y | Y | Y | Y | Y | Y | Y | Y |
| A30 | Yu H, et al | Y | Y | U | Y | Y | Y | Y | Y |
| Q39 | Zeng J, et al | Y | Y | Y | Y | Y | U | Y | Y |
| Q11 | Zhang J, et al | Y | Y | Y | Y | Y | Y | Y | U |
| Q37 | Zhang R, et al | Y | Y | Y | Y | Y | Y | Y | Y |
| Q44 | Zhang W, et al | Y | Y | Y | Y | Y | Y | Y | Y |
| Q10 | Zhao H, et al | Y | Y | Y | Y | Y | Y | Y | Y |
| Q34 | Zhou L, et al | Y | Y | Y | Y | Y | Y | Y | Y |
| Q34* | Yang L, et al | Y | Y | Y | Y | Y | Y | Y | Y |
| Q28 | Zhu C, et al | Y | Y | Y | U | Y | Y | Y | Y |

*: indicates data published in this paper duplicates the data under the same study ID.

**Search terms**

1. (nCoV or n-Cov or 2019-nCoV or "coronavirus disease 2019" or "coronavirus disease-19" or Covid-19 or Covid19 or "novel coronavirus" or COVID or "Middle East Respiratory Syndrome Coronavirus" or "Middle East respiratory syndrome" or MERS or SARS or "severe acute respiratory syndrome" or 2019nCoV or SARS-CoV-2 or Coronavirus or "Corona virus" or corona-virus or "corona viruses" or coronaviruses or SARS-CoV or Orthocoronavirinae or MERS-CoV or "Severe Acute Respiratory Syndrome" or "Middle East Respiratory Syndrome").mp.

2. (secondary attack rate or contact attack rate or close contact or contact transmission or household transmission or contact transmission or contact attack rate or family transmission).mp.

3. ("cluster outbreak*" or "cluster case*" or ((famil* or household) adj4 cluster*)).mp.

4. 2 or 3

5. 1 and 4

6. 5 not (exp animals/ not humans.sh.)

7. limit 6 to last year
